# Long-term low-dose acetylsalicylic use shows protective potential for the development of both vascular dementia and Alzheimer’s disease in patients with coronary heart disease but not in other individuals from the general population: results from two large cohort studies

**DOI:** 10.1101/2021.09.20.21263830

**Authors:** Thi Ngoc Mai Nguyen, Li-Ju Chen, Kira Trares, Hannah Stocker, Bernd Holleczek, Konrad Beyreuther, Hermann Brenner, Ben Schöttker

**Affiliations:** Division of Clinical Epidemiology and Aging Research, German Cancer Research Center (DKFZ), 69120 Heidelberg, Germany; Network Aging Research, University of Heidelberg, 69115 Heidelberg, Germany; Saarland Cancer Registry, 66119 Saarbrücken, Germany

## Abstract

**Objectives:** To investigate the association between long term low-dose acetylsalicylic acid (LDASA) use and the development of all-cause dementia, Alzheimer’s disease (AD) and vascular dementia (VD).

**Design:** Meta-analysis of individual participant data from two prospective cohort studies.

**Setting:** Community-dwelling older adults from Germany (ESTHER) and United Kingdom (UK Biobank).

**Participants:** 5,258 ESTHER and 305,394 UK Biobank participants who were 55 years or older and completed drug assessment were included for analysis.

**Main outcome measures:** Cox regression models with inverse probability of treatment weighting to model the underlying cardiovascular risk were used to assess the associations of LDASA use with all-cause dementia, AD and VD incidence.

**Results:** 476 cases of all-cause dementia, 157 cases of AD and 183 cases of VD were diagnosed over a median of 14.3 years of follow-up in ESTHER. In the UK Biobank, 5,584 participants were diagnosed with all-cause dementia, 2,029 with AD and 1,437 with VD over a median of 11.6 years. The meta-analysis of both cohorts revealed a weak reduction in hazards for all-cause dementia (HR [95% CI]: 0.96 [0.93 to 0.99]). The strongest protective effect of LDASA was observed in participants with coronary heart disease (CHD) in both cohorts, and a significant interaction was detected. In particular, in meta-analysis, a 31% reduction in hazard for AD, 69% for VD and 34% for all-cause dementia were observed (HR [95% CI]: 0.69 [0.59 to 0.80], 0.31 [0.27 to 0.35], 0.46 [0.42 to 0.50], respectively). Furthermore, compared to non-users, users of LDASA for 10 years or longer (who likely use it because they have CHD or a related diagnosis putting them at an increased risk for cardiovascular events) demonstrated a strong protective effect on all dementia outcomes, especially for VD (HR [95% CI]: 0.48 [0.42 to 0.56]) whereas no protective associations were observed with shorter LDASA use.

**Conclusions:** The protective potential of LDASA for all-cause dementia, AD and VD seems to strongly depend on pre-existing CHD and the willingness of patients to take it for a minimum of ten years.

## Introduction

For decades, the non-steroidal anti-inflammatory drug (NSAID) acetylsalicylic acid (ASA) has been widely used at a low dose of 100 to 300 mg per day for secondary prevention of atherosclerotic cardiovascular disease (ASCVD).^1^ Although the risks and benefits of ASA are well known, its potential for neuroprotection is still a matter of debate. Through its anti-inflammatory property, ASA could potentially prevent or delay the onset of Alzheimer’s disease (AD).^2-5^ Moreover, as an anti-thrombotic agent, ASA further helps to reduce cerebrovascular disease, which may also contribute to vascular dementia (VD) prevention.^6^ Systematic reviews and meta-analyses of observational studies but not randomized controlled trials (RCTs) supported the possibility that low-dose acetylsalicylic acid (LDASA) could protect against dementia.^7 8^ However, no population-based, observational cohort study has been performed so far. Studies with long follow-up are especially needed because it has been observed that NSAID use might only protect against AD when initiated long before cognitive decline begins.^9^ Thus, this study investigated the association of LDASA with all-cause dementia, AD, and VD incidence using data from two large, population-based cohorts with more than 10 years of follow-up.

## Methods

### Study design and population

We used data from two prospective cohorts: the ESTHER study from Germany and the UK Biobank from the United Kingdom. ESTHER (full German name: *Epidemiologische Studie zu Chancen der Verhütung, Früherkennung und optimierten Therapie chronischer Erkrankungen in der älteren Bevölkerung*) is an ongoing population-based cohort study whose details have been reported elsewhere.^10^ In brief, 9,940 individuals aged 50-75 years were recruited via their general practitioners (GPs) during a routine health check-up between July 2000 and December 2002. After 2, 5, 8, 11, 14, and 17 years, participants and their GPs were contacted again and asked to complete questionnaires on health status, medical diagnoses and treatments. For this project, we included participants who had a drug assessment at either baseline or 2-year follow-up. After excluding those aged <55 years, with missing dementia diagnosis information from their GPs, or who developed dementia between baseline and the 2-year follow-up, we arrived at N=5,258 for analyses (Supplemental Figure A1).

The UK Biobank is a large-scale, prospective cohort study. Between 2006 and 2010, more than half a million study participants aged 40 to 69 years who lived up to 25 miles from one of 22 study assessment centres in England, Scotland, and Wales were recruited.^11^ At baseline assessment visit, participants completed a touch-screen questionnaire, a brief computer-assisted interview, had physical and functional measurements taken and biological samples collected.^12^ Follow-up of health-related outcomes was enabled through linkage to routinely available data from the UK National Health Service (e.g., mortality, cancer registrations, hospital admissions, primary care data) and in this analysis we used the most up to date available data (up to 31 March 2021 for England and Scotland and 28 February 2018 for Wales). From 502,492 participants at baseline, we excluded those aged <55 years and those with missing information on sex, drug assessment, or dementia diagnosis (Supplemental Figure A2). Overall, 305,394 participants were included.

### Assessment of drugs

In ESTHER, assessment of LDASA use was made by combining the physicians’ questionnaire at baseline (ASA as prescription drug) and the participants’ questionnaire at two-year follow-up (ASA as over-the-counter drug). A participant was considered a LDASA user when there was a record of ASA use at the dose of ≤ 300 mg per day, either at baseline or at two-year follow-up. The cohort entry date was set at the date of arrival of the 2-year follow-up questionnaire.

In the UK Biobank, users of LDASA were identified through the list of codes used by clinic nurses to code drugs that study participants brought to the verbal interview.^13^ The dosage of drugs was not recorded. However, since short-term used drugs and over-the-counter medication were explicitly excluded from this drug assessment, we assumed that ASA was prescribed at a low dose. We further checked this assumption in UK Biobank participants with primary care prescription data whose dosing information was available. Drugs in the GP prescription data were coded using the Read v2, British National Formulary (BNF) and dm+d.^14 15^ Among 221,734 participants of the UK Biobank with drug information available in both interview data and primary care data, 25528 (11.5%) received at least one prescription of LDASA prior to the baseline assessment date and 18454 of these (72.3%) were also identified at the verbal interview. The level of agreement in identifying LDASA users between interview data and primary care data was close to the threshold between moderate and substantial agreement (Cohen’s Kappa coefficient, 0.59, Supplemental Table A1).

### Ascertainment of incident dementia outcomes

In ESTHER, GPs of all participants (including those who had dropped out during follow-up due to illness or death) were asked to provide information on potential dementia diagnoses through standardized questionnaires at the 14- and 17-year follow-up. Diagnoses reported by GPs were confirmed through available medical records of neurologists, psychiatrists, memory or other specialized clinics.^16^ In Germany, diagnosis of AD follows the guidelines of the National Institute on Aging-Alzheimer’s Association workgroups^17^ or the IWG-2 criteria.^18^

In the UK Biobank, incident dementia cases were obtained through algorithmic combinations of linked data from hospital admissions and death registries.^19^ In this analysis, we excluded participants who had already been diagnosed with dementia at study entry, either in hospital admission data or self-reported during the baseline interview. In the ESTHER study, mental inability to fill self-administered questionnaires was an exclusion criterion applied by the GPs during recruitment, which practically excluded individuals with dementia from taking part in the baseline assessment of the cohort.

### Assessment of covariates

In ESTHER, study participants completed a standardized, comprehensive, self-administered questionnaire at baseline, providing information on sociodemographic characteristics, medical history, health status, family history of diseases, and lifestyle factors. Their GPs completed a standardized health check-up form and documented current drug prescriptions. At 2-year follow-up, an additional self-reported questionnaire on medication use was sent to participants. Total and high-density lipoprotein (HDL) cholesterol were assessed in blood samples taken at baseline by enzymatic colourimetric tests (analytes Chol2 2100 and HDLC3 450). C-reactive protein (CRP) was determined by immunoturbidimetry (analyte CRPL3), and serum creatinine was measured by the kinetic Jaffé method (analyte CREJ2) on a Cobas 8000 C701.^20^ The apolipoprotein E (*APOE*) epsilon alleles were determined based on the allelic combination of the single nucleotide polymorphisms (SNPs) *rs7412* and *rs429358* using TaqMan SNP genotyping assays with genotypes analyzed in an endpoint allelic discrimination read using a PRISM 7000 Sequence detection system (Applied Biosystems, Foster City, CA).^21^

In the UK Biobank, participants completed a touch-screen questionnaire at the assessment centre visit, from which socio-demographics (e.g., education, household number, and income), lifestyle (e.g., smoking status, alcohol consumption, and physical activities), psychosocial factors, mental health, and medical history were obtained.^22^ Thereafter, participants interviewed a trained nurse to give further detailed information on major illnesses and disabilities, operations, and regular prescription medication taken. Physical measurements, including blood pressure and anthropometry, and biological samples were also taken.^23^ Total cholesterol and serum creatinine were analyzed by enzymatic tests, CRP by immuno-turbidimetric test, HDL by enzyme immuno-inhibition test, and low-density lipoprotein (LDL) by enzymatic selective protection test utilizing the Beckman Coulter AU5800 system (Beckman Coulter, UK).^24^ Genome-wide genetic data were available for 488377 participants, of whom 49950 were genotyped on the UK BiLEVE Axiom array while the remaining were run on the UK Biobank Axiom array.^25^ As mentioned for ESTHER, depending on the combination of alleles at *rs429358* and *rs7412* variants, individuals were classified according to one of the six common *APOE* genotypes (ε2ε2, ε2ε3, ε3ε3, ε2ε4, ε3ε4 and ε4ε4).

### Statistical analysis

Cox proportional hazards regression models were used to assess the longitudinal associations of LDASA with all-cause dementia, AD, and VD in comparison with study participants using no LDASA. In a simple model, we adjusted for important risk factors for dementia: age, sex, education, *APOE* ⍰*4* genotypes, body mass index (BMI), smoking status, alcohol consumption, physical activity, diabetes, hypertension, coronary heart disease (CHD) and depression. In the main model, we applied the inverse probability of treatment weighting (IPTW) using propensity scores (PS). The results of the logistic regression models used to derive the PS from 57 variables for ESTHER and 47 variables for UK BIOBANK are reported in Supplementary Table A3 and A4. Factors included in the PS are cardiovascular risk or preventive factors (selected based on knowledge of the scientific literature). Weights were assigned to participants based on the inverse of their probability of receiving LDASA, as estimated by the PS. Weights that exceeded the 99^th^ percentiles were set to that threshold.^26 27^ In sensitivity analysis, we also conducted all analyses with PS matching (1:1). The PS matching analysis confirmed our results obtained with IPTW, but resulted in less precise effect estimation; as only about three-quarters of LDASA users could be matched to one control with similar propensity to get the drug prescribed (data not shown).

The IPTW analyses were carried out for the total population and stratified by age, sex, CHD status and *APOE* genotype. Furthermore, interaction tests between stratifying variables and LDASA use were carried out. In a sensitivity analysis, we performed a 5-year lag-time model in which dementia cases in the first 5 years of follow-up were excluded. Finally, we carried out a multivariable logistic regression with restriction to only participants with primary care data available from the UK Biobank because the duration of use was only available from this data source. LDASA users in this analysis were categorized into four groups: non-users, users of ≤5 years, users from 5 to ≤10 years, users of >10 years, depending on the length of LDASA use from the first prescription identifiable in the primary care data to defined end of follow-up: dementia diagnosis, loss to follow-up, death or censoring date (31 March 2021 for study participants from England and Scotland and 28 February 2018 for study participants from Wales).

All statistical analyses were carried out with SAS v.9.4 (North Carolina, USA). All tests were performed two-sided using an α-level of 0.05. To our knowledge, missing values of covariates were missing at random. Multiple imputation of five data sets was undertaken to deal with missing values, and the results of these five imputed datasets were combined by the SAS procedure PROC MIANALYZE. Five imputed datasets have been suggested to be sufficient to get a reasonably accurate estimate.^28^ The proportion of missing values imputed per covariate is shown in Supplemental Table A2. All analyses were first carried out separately in both cohorts and pooled by fixed effects meta-analyses thereafter. Meta-analyses were conducted with the Comprehensive Meta-analysis 2.0 software (Biostat, Englewood, New Jersey, USA).

### Patient and public involvement

These analyses are based on existing data. Patients were not directly engaged in designing the present research question or the outcome measures, nor were they involved in developing plans for recruitment, design, or implementation of the study. Research at the German Cancer Research Center (DKFZ) is, however, generally informed by a Patient Advisory Committee. Results from UK Biobank are routinely disseminated to study participants via the study website and social media outlets.

## Results

Table 1 shows the baseline characteristics of the included study participants of both cohorts. While the UK Biobank study sample included proportionally more study participants younger than 64 years, the proportion of individuals aged above 70 years was higher in the ESTHER cohort (22.0% vs 0.8%). Both cohorts included 46% males. Presumably, due to the age difference, the ESTHER study participants were less physically active, had more frequently HDL levels <40 mg/dL, CRP ≥3 mg/L, and had a higher prevalence of depression, hypertension, diabetes, and CHD. ESTHER’s study participants also smoked more but consumed alcohol less often, and as opposed to 44.1% in UK Biobank, only 11.6% finished 12 years or more of school education. However, this can be explained by the earlier school enrolment in the UK. The distributions of household size, BMI, total cholesterol and *APOE* genotypes were comparable between the two cohorts. The prevalence of LDASA use was also similar: 18.3% in ESTHER vs 18.7% in UK Biobank. In logistic regression models adjusted for age and sex, common factors associated with LDASA use in both cohorts were age, male sex, physical inactivity, current smoking, BMI ≥ 30 kg/m^2^, diabetes, hypertension and CHD (Supplemental Tables A2 and A3).

**Table 1.**
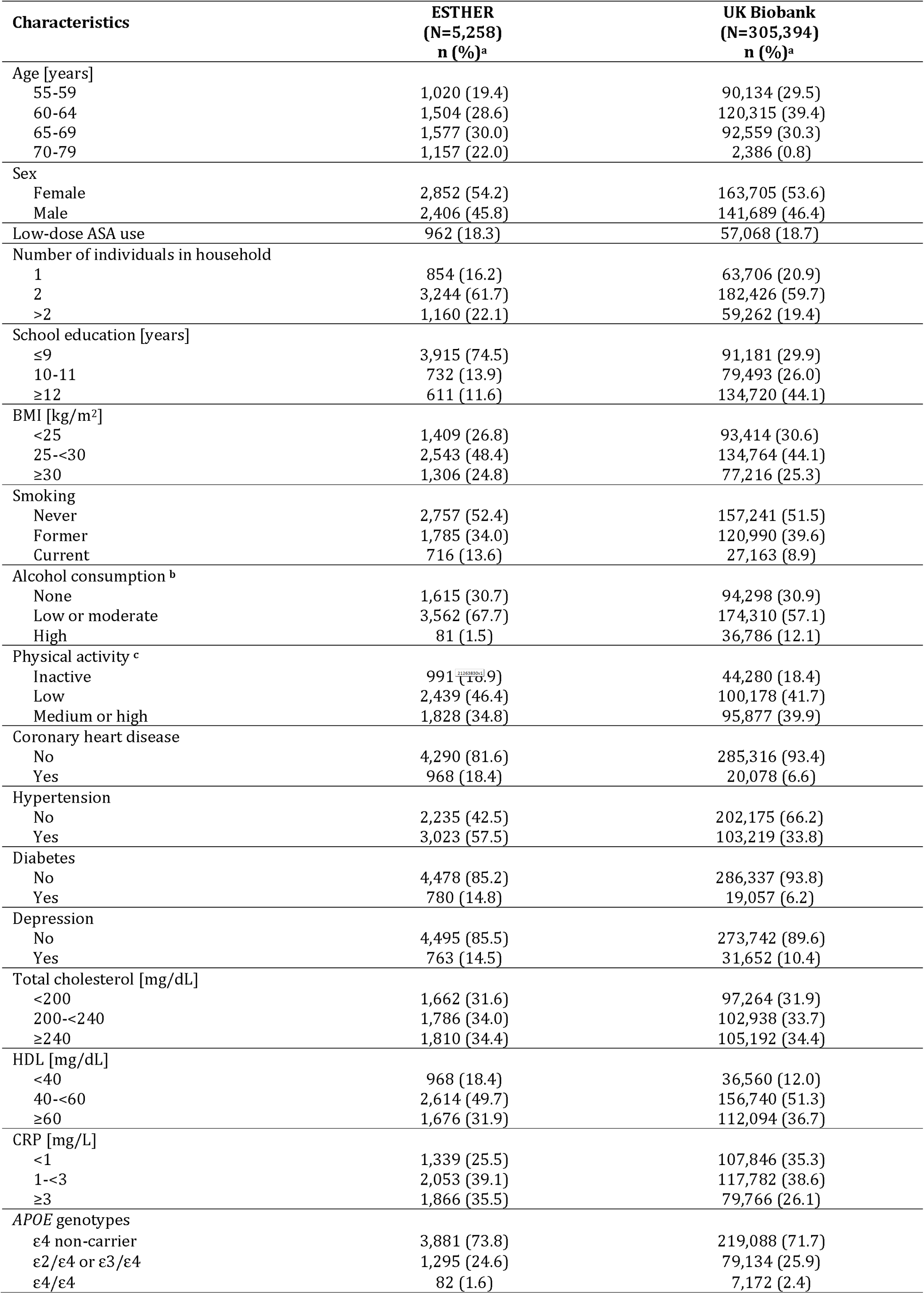
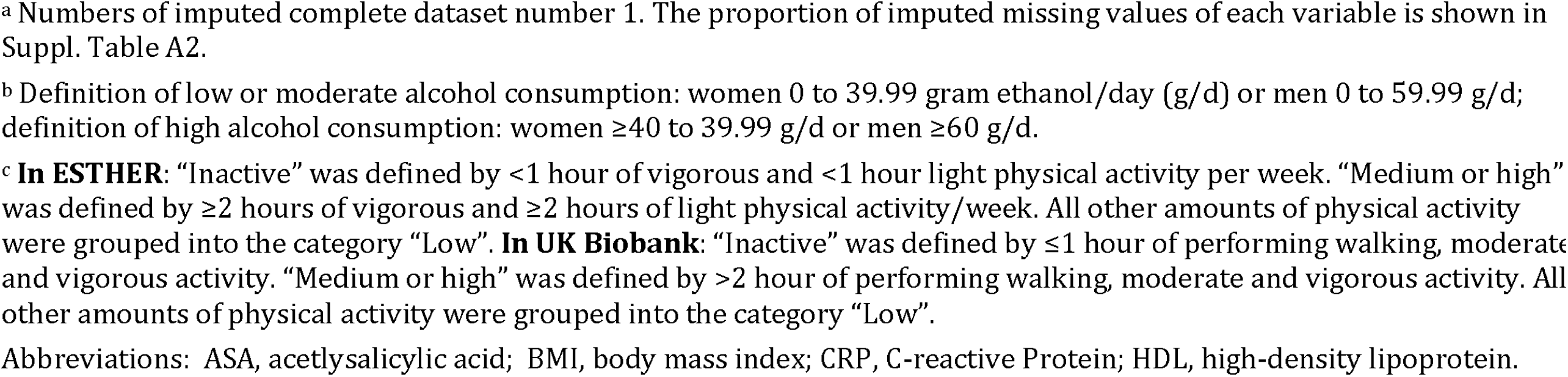
Baseline characteristics of included study participants from the ESTHER (N=5,258) and UK Biobank study (N=305,394)

| Characteristics | ESTHER<br>(N=5,258)<br>n (%)<sup>a</sup> | UK Biobank<br>(N=305,394)<br>n (%)<sup>a</sup> |
| --- | --- | --- |
| Age [years] |  |  |
| 55-59 | 1,020 (19.4) | 90,134 (29.5) |
| 60-64 | 1,504 (28.6) | 120,315 (39.4) |
| 65-69 | 1,577 (30.0) | 92,559 (30.3) |
| 70-79 | 1,157 (22.0) | 2,386 (0.8) |
| Sex |  |  |
| Female | 2,852 (54.2) | 163,705 (53.6) |
| Male | 2,406 (45.8) | 141,689 (46.4) |
| Low-dose ASA use | 962 (18.3) | 57,068 (18.7) |
| Number of individuals in household |  |  |
| 1 | 854 (16.2) | 63,706 (20.9) |
| 2 | 3,244 (61.7) | 182,426 (59.7) |
| >2 | 1,160 (22.1) | 59,262 (19.4) |
| School education [years] |  |  |
| ≤9 | 3,915 (74.5) | 91,181 (29.9) |
| 10-11 | 732 (13.9) | 79,493 (26.0) |
| ≥12 | 611 (11.6) | 134,720 (44.1) |
| BMI [kg/m<sup>2</sup>] |  |  |
| <25 | 1,409 (26.8) | 93,414 (30.6) |
| 25-<30 | 2,543 (48.4) | 134,764 (44.1) |
| ≥30 | 1,306 (24.8) | 77,216 (25.3) |
| Smoking |  |  |
| Never | 2,757 (52.4) | 157,241 (51.5) |
| Former | 1,785 (34.0) | 120,990 (39.6) |
| Current | 716 (13.6) | 27,163 (8.9) |
| Alcohol consumption <sup>b</sup> |  |  |
| None | 1,615 (30.7) | 94,298 (30.9) |
| Low or moderate | 3,562 (67.7) | 174,310 (57.1) |
| High | 81 (1.5) | 36,786 (12.1) |
| Physical activity <sup>c</sup> |  |  |
| Inactive | 991 (18.9) | 44,280 (18.4) |
| Low | 2,439 (46.4) | 100,178 (41.7) |
| Medium or high | 1,828 (34.8) | 95,877 (39.9) |
| Coronary heart disease |  |  |
| No | 4,290 (81.6) | 285,316 (93.4) |
| Yes | 968 (18.4) | 20,078 (6.6) |
| Hypertension |  |  |
| No | 2,235 (42.5) | 202,175 (66.2) |
| Yes | 3,023 (57.5) | 103,219 (33.8) |
| Diabetes |  |  |
| No | 4,478 (85.2) | 286,337 (93.8) |
| Yes | 780 (14.8) | 19,057 (6.2) |
| Depression |  |  |
| No | 4,495 (85.5) | 273,742 (89.6) |
| Yes | 763 (14.5) | 31,652 (10.4) |
| Total cholesterol [mg/dL] |  |  |
| <200 | 1,662 (31.6) | 97,264 (31.9) |
| 200-<240 | 1,786 (34.0) | 102,938 (33.7) |
| ≥240 | 1,810 (34.4) | 105,192 (34.4) |
| HDL [mg/dL] |  |  |
| <40 | 968 (18.4) | 36,560 (12.0) |
| 40-<60 | 2,614 (49.7) | 156,740 (51.3) |
| ≥60 | 1,676 (31.9) | 112,094 (36.7) |
| CRP [mg/L] |  |  |
| <1 | 1,339 (25.5) | 107,846 (35.3) |
| 1-<3 | 2,053 (39.1) | 117,782 (38.6) |
| ≥3 | 1,866 (35.5) | 79,766 (26.1) |
| APOE genotypes |  |  |
| ε4 non-carrier | 3,881 (73.8) | 219,088 (71.7) |
| ε2/ε4 or ε3/ε4 | 1,295 (24.6) | 79,134 (25.9) |
| ε4/ε4 | 82 (1.6) | 7,172 (2.4) |

<sup>a</sup> Numbers of imputed complete dataset number 1. The proportion of imputed missing values of each variable is shown in Suppl. Table A2.
<sup>b</sup> Definition of low or moderate alcohol consumption: women 0 to 39.99 gram ethanol/day (g/d) or men 0 to 59.99 g/d; definition of high alcohol consumption: women $\geq 40$ to 39.99 g/d or men $\geq 60$ g/d.
<sup>c</sup> **In ESTHER:** “Inactive” was defined by <1 hour of vigorous and <1 hour light physical activity per week. “Medium or high” was defined by $\geq 2$ hours of vigorous and $\geq 2$ hours of light physical activity/week. All other amounts of physical activity were grouped into the category “Low”. **In UK Biobank:** “Inactive” was defined by $\leq 1$ hour of performing walking, moderate and vigorous activity. “Medium or high” was defined by >2 hour of performing walking, moderate and vigorous activity. All other amounts of physical activity were grouped into the category “Low”.
Abbreviations: ASA, acetylsalicylic acid; BMI, body mass index; CRP, C-reactive Protein; HDL, high-density lipoprotein.

Among all included n=5,286 participants of the ESTHER study, 476 cases of all-cause dementia were diagnosed during a median follow-up of 14.3 years. Thereof, 157 participants were diagnosed with AD and 182 with VD. Among the included n=305,394 participants from the UK Biobank, 5,584 developed all-cause dementia during a median of 11.6 years follow up, of whom 2,029 were diagnosed with AD and 1,437 with VD. Table 2 shows the longitudinal association between LDASA use and the three dementia outcomes. In the simple model with age, sex, education, *APOE* genotypes, BMI, smoking status, alcohol consumption, physical activity, diabetes, hypertension, CHD and depression as covariates, no significant associations were found between LDASA use and dementia outcomes for ESTHER. On the other hand, LDASA use was found to be significantly associated with increased risk of all-cause dementia and VD (HR [95% CI]: 1.12 [1.05 to 1.20] and 1.27 [1.12 to 1.45], respectively) in the UK Biobank cohort. Results did not change much for ESTHER in the main IPTW model but reversed for UK Biobank: LDASA use was significantly associated with a decreased risk of all-cause dementia (HR [95% CI]: 0.95 [0.92 to 0.99]). When excluding dementia cases that were diagnosed during the first five years of follow-up, an inverse association of LDASA and AD also became statistically significant in ESTHER (HR [95% CI] 0.68 [0.51 to 0.89]). However, results for the UK Biobank did not change much. Meta-analysis of the IPTW model results of both cohorts resulted in weak, inverse associations of LDASA with all dementia outcomes, of which only the pooled effect estimate for all-cause dementia was statistically significant (HR [95% CI]: 0.96 [0.93 to 0.99] (Figure 1).

**Table 2.** Longitudinal association of low-dose ASA use with all-cause and common subtype dementia incidence.

|  | ESTHER, N=5,286 |  |  |  |  |  | UK Biobank, N=305,394 |  |  |  |  |  |
| --- | --- | --- | --- | --- | --- | --- | --- | --- | --- | --- | --- | --- |
|  | All-cause dementia |  | Alzheimer's disease |  | Vascular dementia |  | All-cause dementia |  | Alzheimer's disease |  | Vascular dementia |  |
|  | n<sub>case</sub> | HR<br>(95% CI) | n<sub>case</sub> | HR<br>(95% CI) | n<sub>case</sub> | HR<br>(95% CI) | n<sub>case</sub> | HR<br>(95% CI) | n<sub>case</sub> | HR<br>(95% CI) | n<sub>case</sub> | HR<br>(95% CI) |
| Simple model<sup>a</sup> | 476 | 0.93<br>(0.73, 1.18) | 157 | 0.68<br>(0.43, 1.07) | 182 | 1.00<br>(0.69, 1.45) | 5,584 | **1.12**<br>**(1.05, 1.20)** | 2,029 | 1.11<br>(0.99, 1.25) | 1,437 | **1.27**<br>**(1.12, 1.45)** |
| IPTW model | 476 | 1.01<br>(0.89, 1.14) | 157 | 0.81<br>(0.64, 1.01) | 182 | 1.19<br>(0.97, 1.46) | 5,584 | **0.95**<br>**(0.92, 0.99)** | 2,029 | 1.00<br>(0.94, 1.06) | 1,437 | 0.94<br>(0.88, 1.00) |
| IPTW model plus 5-year lag-time<sup>b</sup> | 409 | 0.93<br>(0.81, 1.08) | 129 | **0.68**<br>**(0.51, 0.89)** | 158 | 1.23<br>(0.99, 1.53) | 4,855 | **0.94**<br>**(0.90, 0.97)** | 1,744 | 1.00<br>(0.94, 1.07) | 1,225 | 0.95<br>(0.88, 1.02) |

<sup>a</sup> Simple model was adjusted for age, sex, education, APOE genotypes, BMI, smoking status, alcohol consumption, physical activity, diabetes, hypertension, coronary heart disease and depression
<sup>b</sup> Dementia cases that happened within the first five years of follow-up were excluded, resulting in a total sample size of N=5,191
Abbreviations: IPTW: Inverse probability of treatment weighting
Note: Statistically significant results are printed in bold

**Figure 1.**
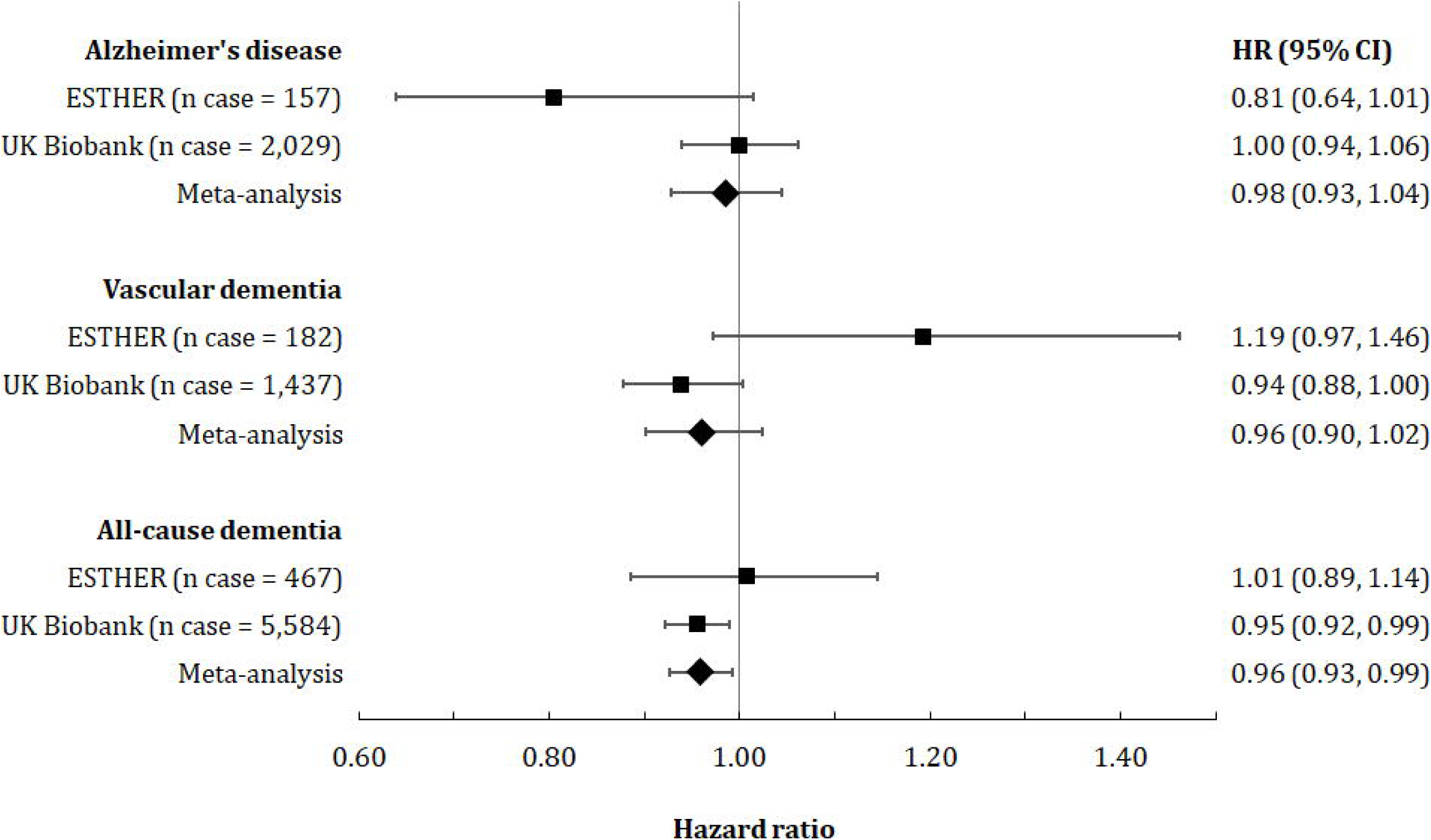
Association between low-dose ASA use and dementia incidence in ESTHER, UK Biobank and the meta-analysis of the two cohorts. The inverse probability of treatment weighting (IPTW) method was used to obtain hazard ratios

Results of the main IPTW model stratified by age, sex, CHD and *APOE* genotype are shown in Supplemental Table A4 and Figure 2. In the meta-analysis, LDASA use was associated with a decreased hazard for all dementia outcomes in participants aged ≥65 years. Furthermore, LDASA use was associated with decreased risk for all-cause dementia and VD incidence in males and not in females. The results did not differ between *APOE ε4* carriers and non-carriers. The strongest protective association of LDASA use was observed in participants with CHD. In particular, LDASA use was associated with 31%, 69% and 54% reduced the risk of developing AD, VD and all-cause dementia, respectively (Figure 2). A statistically significant interaction between CHD and LDASA use was observed for all three outcomes in both cohorts (Supplemental Figure A3). The meta-analyzed p-values for the interaction terms were all <0.001 for all-cause dementia, vascular dementia and Alzheimer’s disease. We also conducted analyses stratified further by both age and CHD (Supplemental Table A6) and by both sex and CHD (Supplemental Table A7). These analyses revealed that CHD was the main effect-modifier in the association between LDASA and dementia outcomes, while age and sex played no important role.

**Figure 2.**
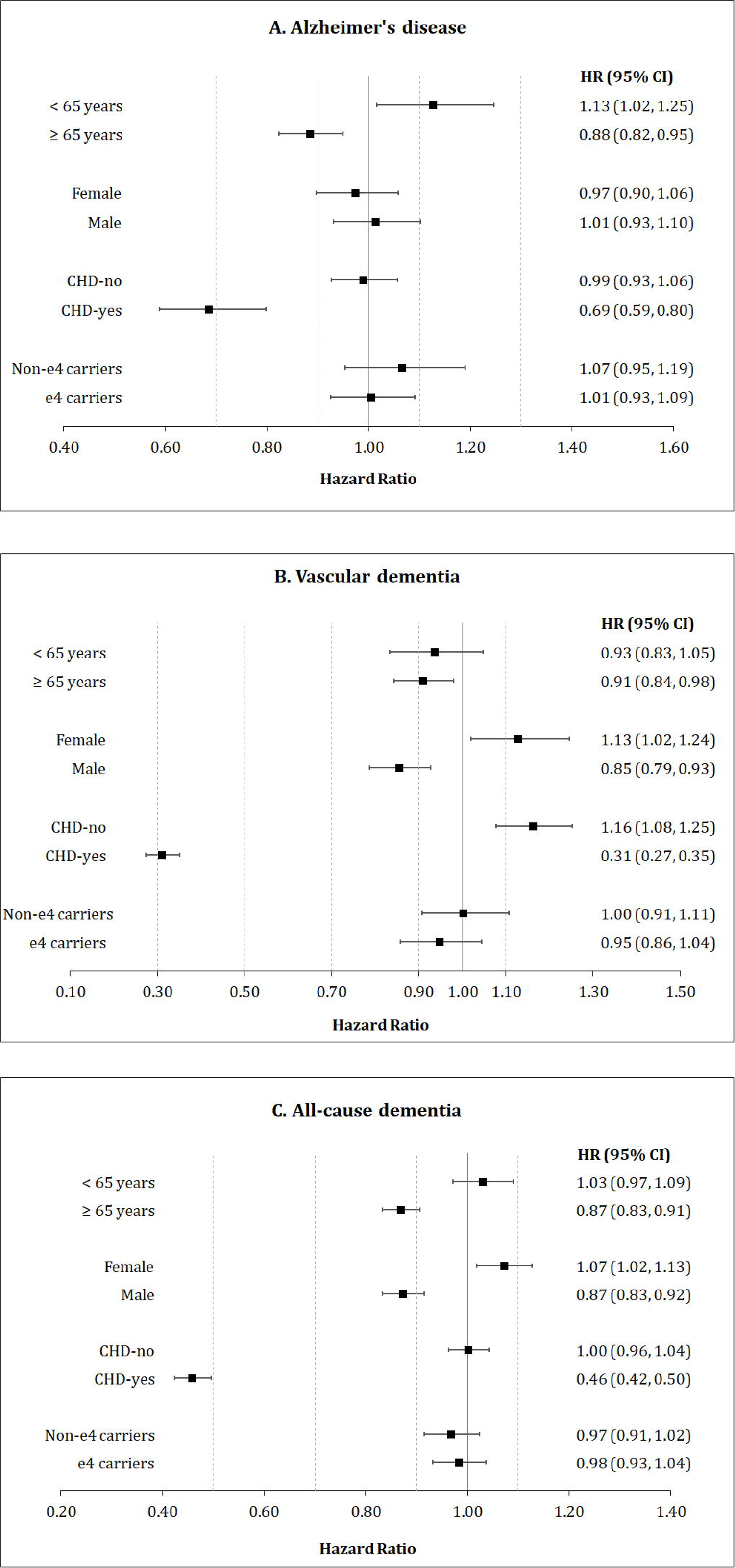
Meta-analysis of the association between low-dose ASA use and dementia outcomes, stratified by age, sex, CHD and *APOE* ε4-carrier status. The inverse probability of treatment weighting (IPTW) method was used to obtain hazard ratios

We included N=136,589 participants aged 55 years and older in the sensitivity analysis on different durations of LDASA use (Table 3). Of those, 100,252 (73.5%) had no LDASA prescription. Two-thirds of LDASA users (N=24,200, 66.6%) had their first LDASA prescription more than 10 years prior to baseline and 36.8% of these long-term users of LDASA had diagnosed CHD. Compared to non-users, those long-term users had approximately half the risk for the three dementia outcomes of non-LDASA users (HR point estimates between 0.48 and 0.58), while users of 5 to 10 years had no decreased risk.

**Table 3.** Association between different duration of low-dose ASA (LDASA) use identified by primary care data (UK Biobank) with all-cause and common subtype dementia incidence (N=136,589)

|  | N total | Prevalence of CHD (%) | All-cause dementia |  | Alzheimer's disease |  | Vascular dementia |  |
| --- | --- | --- | --- | --- | --- | --- | --- | --- |
|  |  |  | n<sub>case</sub> | OR (95% CI)<sup>a</sup> | n<sub>case</sub> | OR (95% CI)<sup>a</sup> | n<sub>case</sub> | OR (95% CI)<sup>a</sup> |
| Non-user | 100,252 | 2.7 | 1,427 | Ref | 521 | Ref | 318 | Ref |
| User of ≤5 years | 2,385 | 32.8 | 216 | **Excluded<sup>b</sup>** | 64 | **Excluded<sup>b</sup>** | 99 | **Excluded<sup>b</sup>** |
| User of >5 to ≤10 years | 9,752 | 36.1 | 309 | 0.95 (0.86, 1.05) | 110 | 1.02 (0.86, 1.20) | 92 | 0.87 (0.73, 1.03) |
| User of > 10 years | 24,200 | 36.8 | 595 | **0.51 (0.47, 0.56)** | 198 | **0.58 (0.51, 0.68)** | 197 | **0.48 (0.42, 0.56)** |

<sup>a</sup> Results of multivariable logistic regression models, adjusted for covariates related to cardiovascular risk (all variables shown in Supplementary Table A3)
<sup>b</sup> Excluded due to protopathic bias (see discussion, strengths and limitations of this study)
Note: Statistically significant results are printed in bold

## Discussion

In this individual participant data meta-analysis of two large cohort studies, the use of LDASA was weakly associated with decreased all-cause dementia incidence but not with AD and VD incidence. However, after stratifying by CHD, it became apparent that only subjects with pre-existing CHD benefited strongly from LDASA use. The results were concordant across both cohorts and validated by significant interaction terms of CHD and LDASA use observed in both cohorts. In addition, when participants were compared based on the length of LDASA use, a strong protective effect was only observed among LDASA users, who started the use 10 years or more prior to baseline. In the latter analysis, 36.8% of users of LDASA for 10 or more years were CHD patients, but we assume that this number is underestimated in the primary care data of the UK Biobank and that all of these long-term LDASA users use it for cardiovascular risk prevention and have CHD or a diagnosis of a related cardiovascular disease. Thus, we do not think that this result contradicts the previous finding that the effectiveness of LDASA strongly depends on pre-existing CHD and that other persons from the general population had no decreased risk of dementia if they were LDASA users.

### Biological mechanisms

Several suggested mechanisms could explain a potential protective effect of ASA use on both AD and VD development. The primary pharmacological activity of ASA is the inhibition of the cyclooxygenase (COX) enzymes, leading to a reduction in the levels of prostaglandins, prostacyclin, and thromboxanes. Those are important in AD pathogenesis,^3 5^ and in the prevention of ischemic brain damage,^29^ a strong risk factor for VD.^6^ Along with the thromboxane pathway, LDASA also inhibits platelet activation and aggregation, which can prevent transient cerebral ischemic attacks and eventually help to enhance the blood flow in the cognitive area.^29 30^

ASA could further reduce the related pathology effect on AD, primarily by reducing amyloid-beta (Aβ). Firstly, it activates the peroxisome-proliferator-activated receptor-γ (PPARγ), which controls the expression of pro-inflammatory genes,^2^ downregulates beta-secretase 1 (BACE1) and thereby reduce amyloid precursor protein (APP) cleavage and Aβ production.^4^ However, it is possible that the 31% reduction in hazard for AD is restricted to those participants with mixed dementia due to the coexistence of AD and VD and thus mediated through a reduction in hazard of the VD component of AD. What speaks in favor of this hypothesis is that neuropathological studies suggest that this type of mixed dementia is a rather common pathological finding in the elderly with a prevalence of about 22%.^31^

Cardio- and cerebrovascular disease and dementia often not only coexist and pose risks for each other, in addition, it is well-known that vascular and neurodegenerative pathologies could interact additionally and synergistically.^32^ Imaging studies have demonstrated that vascular risk factors may contribute to Aβ deposition in the brain.^33 34^ Incident CHD has previously been found to be associated with accelerated long-term cognitive decline.^35 36^ Furthermore, people with pre-existing CHD have an increased risk for recurrent vascular events than CHD-free individuals.^37 38^ Therefore, there are plausible mechanisms in favor of the hypothesis that LDASA is more effective in preventing dementia among CHD patients than in the CHD-free population. Support for this hypothesis can also be drawn from the study of Kern et al.,^39^ which observed that women with high risk of CVD (Framingham risk score of more than 10%) who used LDASA had a decreased loss of cognitive function over a follow-up of 5 years compared to women who did not use LDASA (Mini Mental State Examination (MMSE) score change −0.33 vs −0.95; p=0.028).

### Comparison with other observational studies

Very few longitudinal studies investigated the associations of LDASA use with dementia incidence. None of the existing was population-based, had specifically evaluated the importance of the duration of LDASA use, had VD as an outcome, or tested a potential interaction of ASA use and CHD. A systematic review by H. Li et al. summarized the literature up to April 2020 and we checked in Pubmed that there were no further studies on LDASA use and dementia outcomes as of August 2021.^7^ Overall, a meta-analysis of 8 studies indicated that the use of any dose of ASA did not significantly decrease the risk of developing dementia (pooled relative risk (RR) [95% CI]: 0.94 [0.77 to 1.16]. However, when the authors restricted the meta-analysis to 4 studies with LDASA exposure and the outcome all-cause dementia^39-42^ and 2 studies with LDASA exposure and the outcome AD,^40 41^ LDASA use showed a protective effect against all-cause dementia (pooled RR [95% CI]: 0.82 [0.71 to 0.96]) and AD (pooled RR [95% CI]: 0.54 [0.33 to 0.89]). Among the four studies on all-cause dementia, three reported statistically significant results and only the study of Kern et al.,^39^ which was underpowered (n=41 cases), showed no statistical differences between LDASA users and non-users regarding the 5-year risk of dementia. None of the 4 individual studies is comparable to our study populations from the UK Biobank and ESTHER (general population, age ≥55) because they included either only women,^39^ twins aged 80 years or older,^41^ patients with late-onset depression^42^ or type 2 diabetes patients.^40^

A systematic review of cohort studies on the association of NSAIDs use and AD incidence was conducted by C. Zhang et al. with literature search up to April 2017.^43^ In this review, 16 cohorts with a total of 236022 participants could be included in the meta-analysis and ever use of NSAIDs was also found to be statistically, significantly associated with a reduced risk of AD (RR [95%CI], 0.81 [0.70 to 0.94].^43^ A further meta-analysis limited to 10 studies with ASA (any dose) as exposure yielded a similar RR but without statistical significance (RR [95%CI], 0.89 [0.70 to 1.13]).

In summary, previous observational studies showed some hints that the anti-inflammatory actions of NSAIDs could prevent AD. However, the strongest and most consistent results were observed in LDASA, likely because they are the only group of NSAIDs used mainly long-term for CVD prevention but not occasionally for the indication of pain.

### Findings from randomized controlled trials

The latest systematic review of randomized controlled trials (RCT) on the topic was also performed by H. Li et al. and included two RCTs – both published in 2020.^7^ In the RCT by Matsumoto et al.^44^, 2,536 diabetes patients (age 30–85 years; median: 65 years) without CVD were randomized into receiving LDASA (81 or 100 mg) and were followed over a median of 11.4 years. A tendency towards reducing all-cause dementia risk was observed, but the result was not statistically significant (HR [95%CI]: 0.82 [0.58 to 1.16]). When stratified by sex, however, women in the LDASA group had a lower incidence of dementia compared with those in the non-ASA group (HR [95%CI]: 0.47 [0.25 to 0.86]). The RCT of Ryan et al.^45^ followed 19114 community-dwelling older adults without CVD, aged 70 or older over a median of 4.7 years. The authors observed that there was no difference in the incidence of dementia between the LDASA group (100 mg) and the placebo group (HR [95%CI]: 0.98 [0.83 to 1.15]).

Our results indicate that a protective effect of LDASA on dementia incidence could only be found among CHD patients. This possibly explains the obtained null results in the main analysis of Matsumoto et al. and Ryan et al. in which patients with CVD were excluded. Furthermore, as the average age of the participants was relatively high, and the initiation of LDASA use may have been already too late. Previous research has proposed that the pathological changes of dementia could start more than two decades before the onset of clinical symptoms. In addition, ASA was shown not to affect the cognitive decline in individuals already diagnosed with dementia.^46 47^ In our data, LDASA only showed a protective effect if participants had taken it for at least 10 years, as these people started at a relatively young age and took LDASA sufficiently long. The 5-year treatment period in the study of Ryan et al.^45^ was presumably too short and could be the reason for the null result.

### Public health implications

Potential recommendations for the primary prevention of dementia that could follow our analysis need to be made in context with the existing LDASA guidelines for primary CVD and CRC prevention. A relatively high cardiovascular risk, the initiation of LDASA use at middle age and the willingness of the patient to take LDASA for at least 10 years are the known essential factors for a favourable benefit-risk ratio of LDASA in guidelines for primary prevention of both CVD and colorectal cancer (CRC).^48-61^

A comprehensive decision analysis has concluded that lifetime LDASA use initiated at middle age (40 to 69 years) in persons with higher CVD risk has a favourable benefit-risk ratio in the primary prevention of CVD and colorectal cancer (i.e., outweighs its haemorrhage risks).^60^ Based on this analysis, the United States Preventive Services Task Force (USPSTF) recommends the preventive use of LDASA for all adults aged 50–59⍰years with a 10-year risk of a CVD event greater than 10% and a willingness to take LDASA daily for at least 10 years.^59 61^ Even though the USPSTF guideline targets CVD and CRC prevention, based on our results, following this recommendation could also be an effective measure for the prevention of dementia. Currently, the evidence from RCTs is too weak to include dementia as a factor in decision analyses on LDASA use. However, this might change in the future when more long-term RCTs targeting the right population (middle age and relatively high CVD risk) are published.

### Strengths and limitations of this study

The strengths of this study include the prospective cohort design, the large sample size (n=5258 for ESTHER, n=305394 for UK Biobank), and a long follow-up period (median of 14.3 for ESTHER and 11.6 years for UK Biobank). In addition, evaluating the association of different durations of LDASA use was possible due to utilizing the primary care data. Moreover, by applying the inverse probability of treatment weighting using propensity score, we were able to balance the distribution of CVD risk factors between LDASA users and non-users, and thus, were able to adjust for confounders comprehensively.

Our work inevitably has some shortcomings. First, as with any observational study, residual confounding remains possible, and causation cannot be tested. Protopathic bias was present in the analysis of the primary care data of the UK Biobank with more than 2-fold increased odds ratios for dementia outcomes of LDASA users who initiated LDASA use less than 5 years prior to dementia diagnosis. LDASA use often gets initiated after CVD events and these patients have an increased risk of recurrent CVD events. As most of the dementia diagnoses in the UK Biobank originate from hospital records, an accumulation of first-time dementia diagnoses in the data set among patients hospitalized for recurrent CVD events that happen up to 5 years after LDASA initiation must be expected, since these hospitalizations are needed for awareness of dementia diagnoses in the UK Biobank. Therefore, a sufficient lag time of 5 years between LDASA initiation and dementia diagnosis needed to be ascertained in this analysis by excluding patients with short-term LDASA use. This finding shows that the use of linkage to electronic health records for dementia ascertainment may not be ideal in the UK Biobank, as milder cases of dementia diagnosed in the outpatient setting could have been missed.^62^ The dementia ascertainment in the ESTHER study likely includes more milder dementia cases due to collecting medical records from specialists via participant’s GPs. However, the ESTHER study likewise has some limitations. There was no study protocol for specific dementia diagnostic needed to follow to obtain these diagnoses. The applied procedures in the ESTHER study reflect the current routine done by German clinicians. This can also explain the low proportion of diagnosed AD among the all-cause dementia cases. Many dementia cases have a missing specific diagnosis simply because differential diagnostics are often not made in routine practice in the community setting.

In conclusion, in this analysis of two large, population-based cohort studies from Germany and the UK, LDASA demonstrated a protective potential for AD, VD and all-cause dementia among study participants with pre-existing CHD, but not in other persons from the general population. Furthermore, taking the drug for more than 10 years was critical for detecting the association. This implies that people with CHD may not only profit from long-term LDASA use by reducing their CVD risk but also their dementia risk. The results of this study can only be generalized to mainly Caucasian populations aged 55 years and older, and the findings need to be further tested by RCTs with large sample sizes and long follow-up periods.

## Summary boxes

### What is already known on this topic

□ Neuroinflammation is one of the driving forces in the pathogenesis of Alzheimer’s disease. Cardio- and cerebrovascular conditions are established risk factors for vascular dementia.
□ Usage of acetylsalicylic acid as an anti-thrombotic and anti-inflammatory agent could potentially prevent or delay the onset of dementia.

### What this study adds

□ This is the first long-term population-based study investigating the association between the use of low-dose acetylsalicylic acid (LDASA) and dementia incidence. LDASA demonstrated a protective potential for Alzheimer’s disease, vascular dementia and all-cause dementia among study participants with pre-existing coronary heart disease (CHD), but not in other persons from the general population. Taking the drug for more than 10 years was shown to be essential.
□ In addition, to prevent future cardiovascular events, people with CHD may also benefit from long-term LDASA use by reducing their dementia risk.

## Data Availability

Data from ESTHER is available upon reasonable request that is compatible with participants' informed consent. Data from the UK Biobank (https://www.ukbiobank.ac.uk/) is available to bona fide researchers on application. Part of this reseach was conducted using the UK Biobank Resource under application 69578.

## Acknowledgements

We would like to thank all participants of the ESTHER and UK Biobank cohort as well as the GPs of the ESTHER study and the staff of the UK Biobank assessment centers for their contribution to the studies this research is based on.

## Footnotes

### Contributors

HB designed and led the ESTHER cohort. TNMN and BS generated the idea for the study and formulated the analytical plan. TNMN did the data analyses and drafted the manuscript. BS revised it. All authors contributed valuable intellectual content to the discussion. TNMN and BS are the guarantors of the manuscript and accepts full responsibility for the work and/or the conduct of the study. HB and BS had full access to ESTHER data. TNMN and BS had full access to UK Biobank data used for this study. The corresponding author attests that all listed authors meet authorship criteria and that no others meeting the criteria have been omitted.

### Funding

Data collection of the ESTHER study follow-up used for this project was supported by the Federal Ministry of Education and Research (Berlin, Germany) (grant numbers 01ET0717 and 01GY1320A) and the Saarland Ministry for Social Affairs, Health, Women, and Family Affairs. UK Biobank was established by the Wellcome Trust, Medical Research Council, Department of Health, Scottish government, and Northwest Regional Development Agency. It has also had funding from the Welsh assembly government and the British Heart Foundation. The sponsors had no role in data acquisition or the decision to publish the data.

### No competing interests

All authors have completed the ICMJE uniform disclosure form at www.icmje.org/coi_disclosure.pdf and declare: no support from any organization for the submitted work; no financial relationships with any organizations that might have an interest in the submitted work in the previous three years; no other relationships or activities that could appear to have influenced the submitted work.

### Ethical approval

ESTHER was approved by the Ethics Committee of the Medical Faculty of the University of Heidelberg and Medical Association of Saarland (Application number: 58/2000). UK Biobank received ethical approval from the North West Multicentre Research Ethics Committee (REC reference: 11/NW/03820). Both ESTHER and UK Biobank are conducted in accordance with the 1964 Helsinki declaration and its later amendments.

### Transparancy

TNMN and BS affirm that the manuscript is an honest, accurate, and transparent account of the study being reported; that no important aspects of the study have been omitted; and that any discrepancies from the study as planned (and, if relevant, registered) have been explained.

### Dissemination to participants and related patient and public communities

Findings will be disseminated via the media departments of the authors’ institutes. Results from UK Biobank are routinely disseminated to study participants via the study website and Twitter feed.

### Copyright/license for publication

The Corresponding Author has the right to grant on behalf of all authors and does grant on behalf of all authors, a worldwide licence to the Publishers and its licensees in perpetuity, in all forms, formats and media (whether known now or created in the future), to i) publish, reproduce, distribute, display and store the Contribution, ii) translate the Contribution into other languages, create adaptations, reprints, include within collections and create summaries, extracts and/or, abstracts of the Contribution, iii) create any other derivative work(s) based on the Contribution, iv) to exploit all subsidiary rights in the Contribution, v) the inclusion of electronic links from the Contribution to third party material where-ever it may be located; and, vi) licence any third party to do any or all of the above.

## Notes

### Competing Interest Statement

The authors have declared no competing interest.

