## Supplemental for "Long-term low-dose acetylsalicylic use shows protective potential for the development of both vascular dementia and Alzheimer’s disease in patients with coronary heart disease but not in other individuals from the general population: results from two large cohort studies"

### **Supplemental Figure A1.** Flowchart of study population for ESTHER cohort

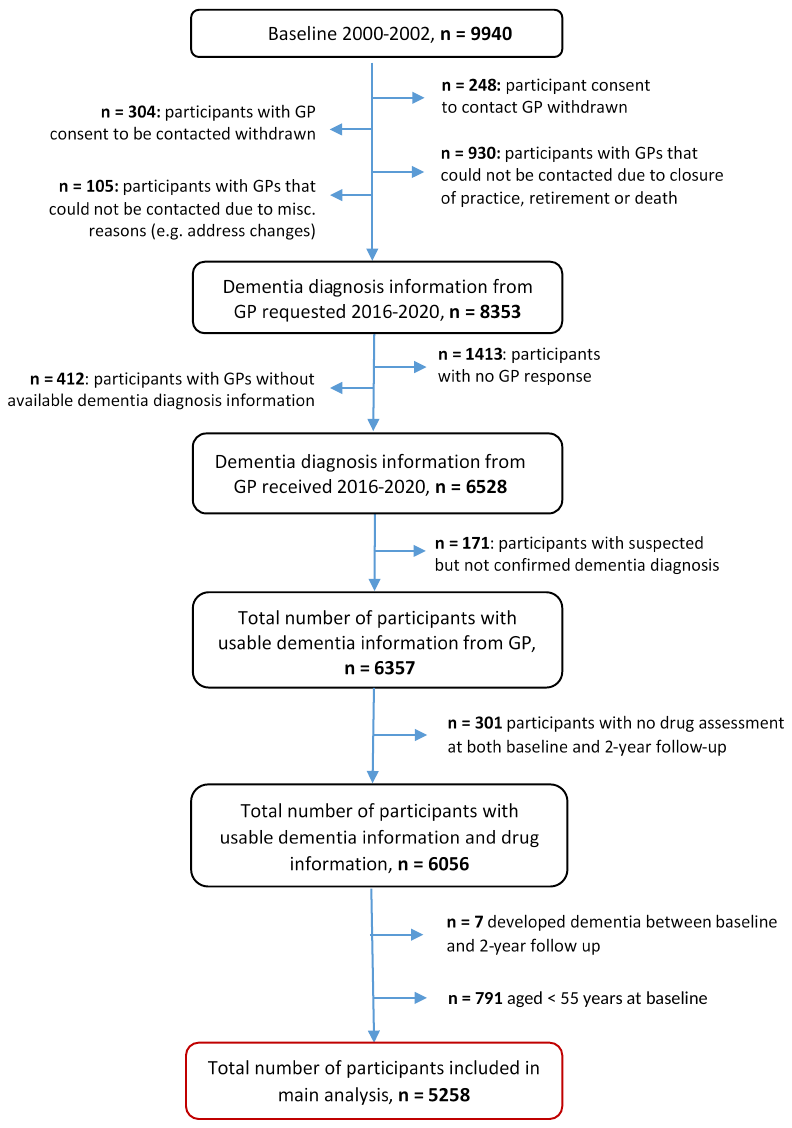

### **Supplemental Figure A2.** Flowchart of study population for UK Biobank cohort

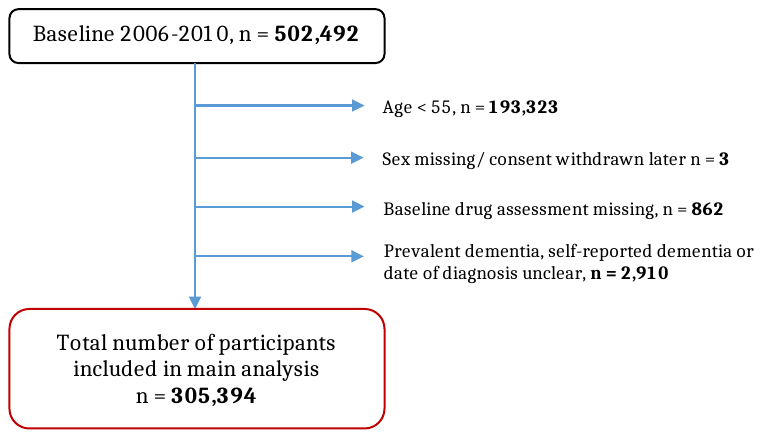

### **Supplemental Figure A3.** Interaction of coronary heart disease and low-dose ASA use for dementia incidence in ESTHER, UK Biobank and the meta-analysis of the two cohorts. The inverse probability of treatment weighting (IPTW) method was used to obtain hazard ratios

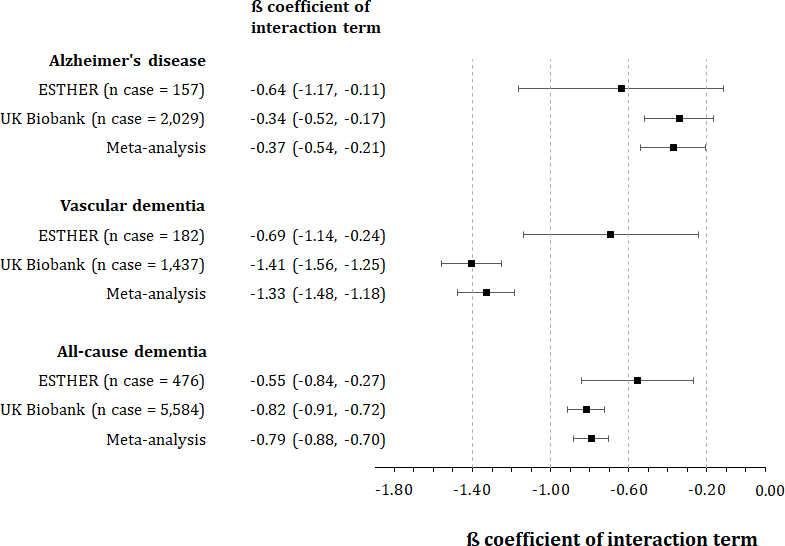

### **Supplemental Table A1.** Cohen’s kappa coefficient demonstrating the agreement in identifying low-dose ASA (LDASA) users between assessment centre interview data and primary care data

|  |  | **Primary care data** | | **Total** |
| --- | --- | --- | --- | --- |
|  |  | **Non-user (n)** | **LDASA user (n)** |  |
| **Interview data at  assessment center** | **Non-user (n)** | 182,726 | 7,074 | 189,800 |
|  | **LDASA user (n)** | 13,480 | 18,454 | 31,934 |
| **Total** |  | 196,206 | 25,528 | **221,734** |

Note: Cohen’s Kappa coefficient (95% CI): 0.59 (0.58, 0.59)

### **Supplemental Table A2.** Proportion of missing values in baseline characteristics of included study participants from the ESTHER (N=5,258) and UK Biobank study (N=305,394) before imputation

| Characteristics | ESTHER  (N=5,258) | UK Biobank  (N=305,394) |
| --- | --- | --- |
|  | **n missing (%)** | **n missing (%)** |
| Age [years] | 0 (0.0) | 0 (0.0) |
| Sex | 0 (0.0) | 0 (0.0) |
| Low-dose ASA use | 0 (0.0) | 0 (0.0) |
| Number of individuals in household | 144 (2.7) | 3,024 (1.0) |
| School education [years] | 129 (2.5) | 5,740 (1.9) |
| BMI [kg/m^2^] | 9 (0.2) | 1,866 (0.6) |
| Smoking | 121 (2.3) | 641 (0.2) |
| Alcohol consumption | 453 (8.6) | 0 (0.0) |
| Physical activity | 12 (0.2) | 65,224 (21.4) |
| Coronary heart disease | 0 (0.0) | 495 (0.2) |
| Hypertension | 0 (0.0) | 462 (0.2) |
| Diabetes | 79 (1.5) | 493 (0.2) |
| Depression | 2 (0.0) | 478 (0.2) |
| Total cholesterol [mg/dL] | 31 (0.6) | 19,880 (6.5) |
| HDL [mg/dL] | 1,910 (36.3) | 43,725 (14.3) |
| CRP [mg/L] | 106 (2.0) | 20,493 (6.7) |
| *APOE* genotypes | 524 (10.0) | 55,235 (18.1) |

### **Supplemental Table A3.** Results of logistic regression models with low-dose acetylsalicylic use as dependent variables used to code propensity scores for ESTHER cohort

| Independent variables | | Age and sex adjusted  OR (95% CI) | Fully adjusted  OR (95% CI) |
| --- | --- | --- | --- |
| Age (per years) | | **1.07 (1.06, 1.09)** | **1.04 (1.02, 1.06)** |
| Sex (male) | | **1.40 (1.21, 1.61)** | 0.99 (0.89, 1.11) |
| School education | |  |  |
|  | ≤ 9 years | Ref | Ref |
|  | 10 to 11 years | 0.93 (0.76, 1.15) | 1.16 (0.91, 1.47) |
|  | ≥ 12 years | 0.82 (0.65, 1.04) | 1.02 (0.78, 1.34) |
| Smoking status | |  |  |
|  | Never | Ref | Ref |
|  | Former | 1.15 (0.98, 1.34) | 1.13 (0.92, 1.37) |
|  | Current | **1.35 (1.10, 1.66)** | **1.57 (1.22, 2.02)** |
| Alcohol consumption ^a^ | |  |  |
|  | None | Ref | Ref |
|  | Low or moderate | **0.84 (0.72, 0.99)** | 1.09 (0.89, 1.32) |
|  | High | 0.64 (0.32, 1.25) | 1.01 (0.49, 2.09) |
| Physical activities ^b^ | |  |  |
|  | Medium or high | Ref | Ref |
|  | Low | **1.20 (1.02, 1.42)** | 1.01 (0.84, 1.21) |
|  | Inactive | **1.42 (1.15, 1.74)** | **1.04 (1.02, 1.06)** |
| BMI (kg/m^2^) | |  |  |
|  | <25 | Ref | Ref |
|  | 25-<30 | 0.92 (0.80, 1.06) | 0.99 (0.80, 1.22) |
|  | ≥30 | **1.40 (1.20, 1.64)** | 1.03 (0.80, 1.33) |
| Number of household  members (per person) | | 1.04 (0.95, 1.13) | 1.07 (0.97, 1.19) |
| Regular use of NSAIDs | | 0.67 (0.40, 1.13) | **0.52 (0.29, 0.95)** |
| *APOE* genotype | |  |  |
|  | ε2/ε2 | Ref | Ref |
|  | ε2/ε3 | 0.93 (0.73, 1.19) | 0.84 (0.32, 2.26) |
|  | ε2/ε4 | 1.38 (0.96, 1.98) | 1.33 (0.48, 3.67) |
|  | ε3/ε3 | 0.94 (0.80, 1.11) | 0.86 (0.32, 2.29) |
|  | ε3/ε4 | 1.04 (0.85, 1.25) | 0.85 (0.32, 2.24) |
|  | ε4/ε4 | 1.31 (0.68, 2.53) | 0.91 (0.27, 3.09) |
| Limit in moderate activities | |  |  |
|  | No | Ref | Ref |
|  | To certain extent | **1.52 (1.31, 1.76)** | 1.01 (0.82, 1.26) |
|  | Severe limitation | **1.39 (1.08, 1.79)** | 0.78 (0.53, 1.16) |
| Limit in climbing staircase | |  |  |
|  | No | Ref | Ref |
|  | To certain extent | **1.43 (1.23, 1.65)** | **1.37 (1.11, 1.69)** |
|  | Servere limitation | **1.67 (1.38, 2.02)** | 1.26 (0.92, 1.73) |
| Limit in the amount of work  or activities of daily living  due to physical impairments | | **1.66 (1.44, 1.92)** | 1.00 (0.76, 1.32) |
| Limit in the types of work or  activities of daily living due  to physical impairments | | **1.69 (1.45, 1.96)** | 1.18 (0.88, 1.57) |
| Limit in the amount of work  or activities of daily living  due to psychological problems | | **1.40 (1.19, 1.64)** | 1.09 (0.77, 1.55) |
| Limit in the types of work or  activities of daily living due  to psychological problems | | **1.36 (1.16, 1.61)** | 0.95 (0.67, 1.36) |
| Feeling calm and relaxing | |  |  |
|  | Always | Ref | Ref |
|  | Sometimes | **1.31 (1.12, 1.52)** | 1.22 (0.99, 1.51) |
|  | Rarely | 1.28 (0.99, 1.65) | 1.09 (0.76, 1.58) |
| Feeling full of energy | |  |  |
|  | Always | Ref | Ref |
|  | Sometimes | 1.00 (0.86, 1.16) | 0.81 (0.65, 1.00) |
|  | Rarely | **1.53 (1.27, 1.85)** | 0.94 (0.68, 1.30) |
| Feeling sad or discourage | |  |  |
|  | Rarely | Ref | Ref |
|  | Sometimes | **1.29 (1.11, 1.51)** | 1.01 (0.81, 1.25) |
|  | Always | 1.24 (0.85, 1.78) | 1.1 (0.69, 1.76) |
| Limit in social activities due  to physical or psychological  problems | |  |  |
|  | Rarely | Ref | Ref |
|  | Sometimes | **1.36 (1.14, 1.61)** | 0.99 (0.78, 1.26) |
|  | Always | 1.12 (0.83, 1.51) | 0.67 (0.45, 1.01) |
| Estimated glomerular  filtration rate  (per 10 mL/min/1.73 m^2^) | | **0.95 (0.91, 0.98)** | 0.97 (0.93, 1.01) |
| Total cholesterol  (per 10 mg/dL) | | **0.98 (0.97, 0.99)** | 1.00 (0.99, 1.02) |
| High-density lipoprotein  (per 10 mg/dL) | | **0.87 (0.82, 0.93)** | 0.98 (0.92, 1.04) |
| C-reactive protein (mg/L) | |  |  |
|  | <1 | Ref | Ref |
|  | 1-<3 | 1.09 (0.94, 1.26) | 0.99 (0.80, 1.22) |
|  | ≥3 | 1.04 (0.90, 1.21) | 0.86 (0.68, 1.08) |
| Use of heart glycoside | | **1.84 (1.24, 2.75)** | 0.87 (0.53, 1.43) |
| Use of angiotensin receptor  blockers | | **1.58 (1.21, 2.08)** | 1.11 (0.81, 1.52) |
| Use of angiotensin-converting- enzyme inhibitors | | **2.33 (1.98, 2.74)** | **1.30 (1.06, 1.6)** |
| Use of insulin | | **3.01 (2.08, 4.36)** | 1.38 (0.86, 2.23) |
| Use of oral diabetes medications | | **1.75 (1.34, 2.29)** | 1.00 (0.69, 1.46) |
| Use of lipid lowering medications | | **3.74 (3.12, 4.49)** | 1.45 (0.92, 2.26) |
| Use of osteoporosis medications | | 0.63 (0.28, 1.42) | 0.66 (0.28, 1.56) |
| Use of statins | | **4.33 (3.55, 5.29)** | 1.28 (0.79, 2.08) |
| Use of vasodialators | | **6.52 (5.13, 8.29)** | **1.71 (1.25, 2.32)** |
| Angina pectoris/ coronary heart  disease (CHD) | | **5.17 (4.34, 6.15)** | **2.05 (1.63, 2.59)** |
| Impaired blood flow in the legs | | **1.78 (1.50, 2.11)** | **1.41 (1.15, 1.72)** |
| Bypass operation | | **10.22 (6.88, 15.17)** | **2.05 (1.27, 3.29)** |
| Cholezystectomy | | **1.34 (1.09, 1.65)** | 1.16 (0.85, 1.57) |
| Depression | | **1.24 (1.01, 1.51)** | 0.96 (0.75, 1.23) |
| Neurodermatitis | | 1.23 (0.87, 1.74) | 1.24 (0.84, 1.84) |
| Dilatation of the carotis | | **9.83 (3.86, 25.05)** | **4.15 (1.41, 12.21)** |
| Dilatation of coronary vessels | | **9.04 (7.08, 11.55)** | **2.55 (1.85, 3.53)** |
| Gout | | **1.39 (1.12, 1.72)** | 1.17 (0.91, 1.49) |
| Glaucoma | | **1.36 (1.00, 1.85)** | 1.27 (0.89, 1.8) |
| Gall stones | | **1.30 (1.09, 1.55)** | 1.01 (0.78, 1.31) |
| Infection with helicobacter  pylori | | 0.88 (0.67, 1.17) | **0.71 (0.51, 0.99)** |
| Heart failure | | **2.38 (1.97, 2.89)** | 0.85 (0.66, 1.09) |
| Hypertension | | **1.87 (1.57, 2.23)** | **1.66 (1.36, 2.04)** |
| Cataract | | **1.45 (1.18, 1.78)** | 1.20 (0.95, 1.52) |
| Stroke | | **4.41 (3.18, 6.12)** | **3.06 (2.12, 4.42)** |
| Cancer (all causes) | | 1.15 (0.89, 1.48) | 1.26 (0.95, 1.68) |
| Diabetes mellitus type 2 | | **1.87 (1.57, 2.23)** | 1.17 (0.89, 1.54) |
| Dyslipidemia | | **1.68 (1.46, 1.94)** | 1.00 (0.83, 1.21) |
| Myocardial infarction | | **6.70 (5.22, 8.61)** | 1.39 (0.99, 1.97) |
| Endoprothesis hip | | 0.80 (0.46, 1.41) | 0.89 (0.48, 1.63) |
| Endoprothesis knee | | 0.52 (0.20, 1.37) | 0.47 (0.16, 1.33) |
| Femoral neck fracture | | 1.00 (0.46, 2.17) | 0.68 (0.27, 1.7) |
| Ulcer | | 1.07 (0.86, 1.32) | 1.03 (0.80, 1.33) |

^a^ Definition of low or moderate alcohol consumption: women 0 to 39.99 gram ethanol/day (g/d) or men 0 to 59.99 g/d; definition of high alcohol consumption: women ≥40 to 39.99 g/d or men ≥60 g/d.

^b^ “Inactive” was defined by <1 hour of vigorous or <1 hour light physical activity per week. “Medium or high” was defined by ≥2 hours of vigorous and ≥2 hours of light physical activity/week. All other amounts of physical activity were grouped into the category “Low”.

Statistically significant results are in bold.

### **Supplemental Table A4.** Results of logistic regression models with low-dose acetylsalicylic use as dependent variables used to code propensity scores for UK Biobank cohort

| Independent variables | | Age and sex adjusted  OR (95% CI) | Fully adjusted  OR (95% CI) |
| --- | --- | --- | --- |
| Age (per years) | | **1.09 (1.09, 1.10)** | **1.05 (1.04, 1.05)** |
| Sex (male) | | **2.17 (2.13, 2.21)** | **1.22 (1.2, 1.24)** |
| School education | |  |  |
|  | ≤ 9 years | Ref | Ref |
|  | 10 to 11 years | **0.84 (0.82, 0.86)** | 0.99 (0.97, 1.01) |
|  | ≥ 12 years | **0.75 (0.74, 0.77)** | **1.05 (1.03, 1.07)** |
| Smoking status | |  |  |
|  | Never | Ref | Ref |
|  | Former, occasionally | **0.89 (0.87, 0.92)** | 1.00 (0.96, 1.04) |
|  | Former, regularly | **1.33 (1.30, 1.35)** | **1.04 (1.01, 1.07)** |
|  | Current, occasionally | **1.12 (1.05, 1.18)** | **1.19 (1.10, 1.28)** |
|  | Current, regularly | **1.32 (1.27, 1.36)** | **1.35 (1.29, 1.42)** |
| Alcohol consumption ^a^ | |  |  |
|  | None | Ref | Ref |
|  | Low | **0.85 (0.83, 0.86)** | **1.05 (1.02, 1.08)** |
|  | Moderate | **0.88 (0.86, 0.90)** | **1.10 (1.06, 1.14)** |
|  | High | 1.01 (0.98, 1.04) | **1.22 (1.17, 1.27)** |
| Physical activities ^b^ | |  |  |
|  | Medium or high | Ref | Ref |
|  | Low | **1.12 (1.10, 1.15)** | **0.96 (0.94, 0.98)** |
|  | Inactive | **1.39 (1.35, 1.43)** | **1.01 (1.00, 1.03)** |
| BMI (kg/m^2^) | |  |  |
|  | <20 | Ref | Ref |
|  | 20-<25 | **1.12 (1.03, 1.22)** | 0.97 (0.94, 1) |
|  | 25-<30 | **1.61 (1.48, 1.76)** | **1.06 (1.03, 1.09)** |
|  | ≥30 | **2.67 (2.45, 2.92)** | **1.10 (1.06, 1.15)** |
| Frequency of friends/family  visits | |  |  |
|  | Rarely or never | Ref | Ref |
|  | Once per month | **0.92 (0.89, 0.95)** | 0.98 (0.93, 1.03) |
|  | Once per week | **0.94 (0.92, 0.96)** | 1.01 (0.96, 1.05) |
|  | 2-4 times per week | 1.01 (0.99, 1.03) | 1.00 (0.95, 1.05) |
|  | Almost daily | **1.15 (1.11, 1.18)** | 1.01 (0.96, 1.06) |
| Income | |  |  |
|  | Less than 18,000 | Ref | Ref |
|  | 18,000 to 30,000 | **0.96 (0.94, 0.98)** | 1.03 (1.00, 1.07) |
|  | 30,000 to 51,999 | **0.84 (0.82, 0.86)** | **1.06 (1.01, 1.10)** |
|  | 52,000 to 100,000 | **0.80 (0.78, 0.83)** | **1.09 (1.03, 1.16)** |
|  | Greater than 100,000 | **0.92 (0.87, 0.98)** | **1.28 (1.16, 1.40)** |
| Number of household  members (per person) | | **0.97 (0.96, 0.98)** | 0.99 (0.98, 1.00) |
| *APOE* genotype | |  |  |
|  | ε2/ε2 | Ref | Ref |
|  | ε2/ε3 | **0.88 (0.86, 0.91)** | **1.05 (1.01, 1.09)** |
|  | ε2/ε4 | **0.92 (0.86, 0.99)** | **1.07 (1.03, 1.10)** |
|  | ε3/ε3 | 0.99 (0.97, 1.02) | 1.03 (0.86, 1.24) |
|  | ε3/ε4 | **1.08 (1.06, 1.11)** | 0.98 (0.81, 1.18) |
|  | ε4/ε4 | **1.11 (1.03, 1.18)** | 1.04 (0.87, 1.23) |
| Numbers of drug taking (per  medication) | | **1.43 (1.43, 1.44)** | **1.35 (1.34, 1.36)** |
| Forced expiratory volume in  one second (per liter) | | **0.70 (0.69, 0.71)** | **1.03 (1.01, 1.05)** |
| HbA_1C_ (per percent) | | **1.06 (1.06, 1.06)** | 1.00 (1.00, 1.01) |
| Numbers of comorbidiy | | **1.33 (1.32, 1.33)** | **0.90 (0.90, 0.91)** |
| Overall health rating | |  |  |
|  | Excellent | Ref | Ref |
|  | Good | **0.68 (0.67, 0.69)** | **1.15 (1.11, 1.19)** |
|  | Fair | **2.03 (1.99, 2.07)** | **1.12 (1.07, 1.17)** |
|  | Poor | **2.80 (2.70, 2.91)** | **0.74 (0.69, 0.79)** |
| Diabetes | |  |  |
|  | No | **Ref** | **Ref** |
|  | Untreated | **3.42 (3.22, 3.63)** | **1.65 (1.53, 1.78)** |
|  | Treated with oral  medications | **5.96 (5.72, 6.21)** | **1.35 (1.28, 1.44)** |
|  | Treated with insulin | **7.21 (6.73, 7.72)** | **1.42 (1.29, 1.56)** |
| Hypertension | |  |  |
|  | No | Ref | Ref |
|  | Untreated hypertension | **0.79 (0.76, 0.82)** | **1.51 (1.45, 1.58)** |
|  | Treated hypertension | **4.21 (4.13, 4.29)** | **1.48 (1.44, 1.52)** |
| Estimated glomerular  filtration rate (per 10 mL/ min/1.73 m^2^) | | **0.90 (0.89, 0.91)** | **1.02 (1.01, 1.03)** |
| Waist circumference (per 10 cm) | | **1.34 (1.33, 1.35)** | 1.01 (1.00, 1.03) |
| Total cholesterol (per 10 mg/dL) | | **0.83 (0.83, 0.83)** | **0.95 (0.94, 0.96)** |
| High-density lipoprotein  (per 10 mg/dL) | | **0.78 (0.77, 0.79)** | 1.01 (1.00, 1.02) |
| Low-density lipoprotein  (per 10 mg/dL) | | **0.79 (0.78, 0.79)** | **1.02 (1.01, 1.04)** |
| Diastolic blood pressure  (per 10 mmHg) | | **0.88 (0.87, 0.89)** | 0.99 (0.98, 1.00) |
| Systolic blood pressure  (per 10 mmHg) | | **0.98 (0.98, 0.99)** | 1.01 (1.00, 1.01) |
| C-reactive protein (mg/L) | |  |  |
|  | <1 | Ref | Ref |
|  | 1-<3 | **1.03 (1.01, 1.05)** | **0.96 (0.93, 0.99)** |
|  | ≥3 | **1.14 (1.11, 1.17)** | **0.88 (0.85, 0.9)** |
| Taking antidepressants | | **1.49 (1.44, 1.55)** | **0.62 (0.59, 0.65)** |
| Taking lipid lowering  medications | | **9.11 (8.93, 9.31)** | **2.44 (2.36, 2.51)** |
| Frequency of depressed mood  in the past 2 weeks | | **1.53 (1.46, 1.60)** | **1.09 (1.02, 1.15)** |
| Taking NSAIDs for pain relief  purpose | | **0.75 (0.73, 0.77)** | **0.59 (0.57, 0.61)** |
| Anxiety | | 1.07 (0.98, 1.16) | 1.10 (0.99, 1.22) |
| Arthritis | | **1.13 (1.10, 1.16)** | **0.81 (0.78, 0.84)** |
| Asthma | | **1.05 (1.02, 1.08)** | **0.64 (0.62, 0.67)** |
| Bronchitis | | **1.50 (1.41, 1.59)** | **0.84 (0.77, 0.91)** |
| Cancer | | **0.94 (0.91, 0.97)** | **0.95 (0.91, 0.99)** |
| Coronary heart diseases | | **19.93 (19.21, 20.67)** | **0.67 (0.57, 0.78)** |
| Chronic obstructive pulmonary  disease | | **1.32 (1.17, 1.49)** | **6.90 (6.61, 7.21)** |
| History of bipolar and major  depression status | | **1.06 (1.03, 1.09)** | **0.91 (0.87, 0.95)** |
| Fatigue | | 0.94 (0.79, 1.11) | 0.86 (0.70, 1.05) |
| Fractured/broken bones in  last 5 years | | 1.02 (0.99, 1.05) | 0.98 (0.94, 1.02) |
| Gout | | **1.53 (1.45, 1.62)** | **0.66 (0.62, 0.71)** |
| Heart failure | | **3.15 (2.45, 4.04)** | **0.65 (0.48, 0.89)** |
| Kidney failure | | **2.34 (1.69, 3.23)** | 0.76 (0.50, 1.15) |
| Osteoporosis | | 0.95 (0.90, 1.00) | **0.57 (0.54, 0.62)** |
| Parkinson | | 1.00 (0.85, 1.17) | **0.61 (0.50, 0.75)** |
| Peripheral vascular disease | | **2.66 (2.24, 3.17)** | **1.86 (1.49, 2.33)** |
| Stroke | | **9.25 (8.72, 9.82)** | **4.06 (3.79, 4.36)** |
| Frequency of tiredness /  lethargy in last 2 weeks | | **1.62 (1.57, 1.67)** | 0.96 (0.92, 1.00) |

^a^ Definition of low alcohol consumption: women 0 to 19.99 gram ethanol/day (g/d) or men 0 to 39.99 g/d; definition of moderate alcohol consumption: women 20 to 39.99 g/d or men 40 to 59.99 g/d; definition of high alcohol consumption: women ≥40 to 39.99 g/d or men ≥60 g/d.

^b^ “Inactive” was defined by ≤1 hour of performing walking, moderate and vigorous activity. “Medium or high” was defined by >2 hour of performing walking, moderate and vigorous activity. All other amounts of physical activity were grouped into the category “Low”.

Statistically significant results are in bold.

### **Supplemental Table A5.** Longitudinal association between low-dose ASA use with all-cause and common subtype dementia incidence, stratified by age, sex, coronary heart disease and APOE genotype^a^

|  |  | **ESTHER, N=5,286** | | | | | |  | **UK Biobank, N=305,394** | | | | | | |
| --- | --- | --- | --- | --- | --- | --- | --- | --- | --- | --- | --- | --- | --- | --- | --- |
|  |  | **All-cause  dementia** | | **Alzheimer’s  disease** | | **Vascular  dementia** | |  | **All-cause  dementia** | | **Alzheimer’s  disease** | | | **Vascular  dementia** | |
|  |  | **n_case_** | **HR**  **(95% CI)** | **n_case_** | **HR**  **(95% CI)** | **n_case_** | **HR**  **(95% CI)** |  | n_case_ | **HR**  **(95% CI)** | **n_case_** | **HR**  **(95% CI)** | **n_case_** | | **HR**  **(95% CI)** |
| **Age** | 55-64 years | 100 | 1.30  (0.98, 1.73) | 35 | 1.08  (0.66, 1.78) | 35 | 1.31  (0.79, 2.16) |  | 2,100 | 1.02  (0.96, 1.08) | 720 | **1.13**  **(1.02, 1.25)** | 486 | | 0.92  (0.82, 1.03) |
|  | ≥ 65 years | 376 | 0.87  (0.75, 1.00) | 122 | 0.69  (0.53, 0.89) | 147 | 1.06  (0.85, 1.33) |  | 3,484 | **0.87**  **(0.83, 0.91)** | 1,309 | **0.90**  **(0.84, 0.97)** | 951 | | **0.89**  **(0.82, 0.96)** |
| **Sex** | Female | 247 | 1.03  (0.86, 1.23) | 92 | 0.83  (0.62, 1.12) | 93 | 1.09  (0.82, 1.46) |  | 2,593 | **1.08**  **(1.02, 1.13)** | 1,031 | 0.99  (0.91, 1.08) | 592 | | **1.13**  **(1.02, 1.26)** |
|  | Male | 229 | 0.99  (0.82, 1.19) | 65 | 0.75  (0.52, 1.09) | 89 | 1.32  (0.99, 1.75) |  | 2,991 | **0.87**  **(0.82, 0.91)** | 998 | 1.03  (0.94, 1.12) | 845 | | **0.82**  **(0.75, 0.89)** |
| **Coronary  heart  disease** | No | 357 | 1.11  (0.95, 1.3) | 124 | 0.96  (0.74, 1.26) | 133 | **1.31**  **(1.03, 1.66)** |  | 4,738 | 0.99  (0.96, 1.04) | 1,757 | 0.99  (0.93, 1.06) | 1,136 | | **1.15**  **(1.06, 1.24)** |
|  | Yes | 119 | **0.63**  **(0.49, 0.80)** | 33 | **0.51**  **(0.32, 0.80)** | 49 | **0.66**  **(0.45, 0.96)** |  | 846 | **0.44**  **(0.41, 0.48)** | 272 | **0.71**  **(0.60, 0.84)** | 301 | | **0.28**  **(0.25, 0.32)** |
| ***APOE*  genotype** | *APOE*  ε4 non-carriers | 260 | 1.08  (0.91, 1.28) | 70 | 1.29  (0.94, 1.79) | 106 | 1.07  (0.82, 1.4) |  | 2,063 | 0.96  (0.90, 1.01) | 598 | 1.04  (0.92, 1.17) | 554 | | 0.99  (0.89, 1.10) |
|  | *APOE* ε4  carriers | 167 | 0.90  (0.72, 1.12) | 70 | **0.42**  **(0.28, 0.62)** | 55 | 1.36  (0.93, 1.99) |  | 2,452 | 0.99  (0.94, 1.04) | 1,041 | 1.04  (0.96, 1.14) | 606 | | 0.92  (0.83, 1.02) |

^a^ Models used inverse probability of treatment weighting

Statistically significant results are in bold

### **Supplemental Table A6.** Meta-analysis of the association between low-dose ASA use with all-cause and common subtype dementia incidence, stratified by both age and coronary heart disease

| **CHD** | **Age** | **All-cause dementia** | | | **Alzheimer’s disease** | | | **Vascular dementia** | | |
| --- | --- | --- | --- | --- | --- | --- | --- | --- | --- | --- |
|  |  | **n_case_**  **UK Biobank** | **n_case_**  **ESTHER** | **Meta-analysis**  **HR (95% CI)** | **n_case_**  **UK Biobank** | **n_case_**  **ESTHER** | **Meta-analysis**  **HR (95% CI)** | **n_case_**  **UK Biobank** | **n_case_**  **ESTHER** | **Meta-analysis**  **HR (95% CI)** |
| No | 55 – 64 years | 1,814 | 83 | **1.08 (1.01, 1.15)** | 637 | 31 | 1.05 (0.94, 1.18) | 384 | 27 | 1.09 (0.95, 1.25) |
|  | ≥ 65 years | 2,924 | 274 | **0.94 (0.90, 0.99)** | 1,120 | 93 | 0.94 (0.87, 1.02) | 752 | 106 | **1.17 (1.07, 1.28)** |
| Yes | 55 – 64 years | 286 | 17 | **0.39 (0.34, 0.45)** | 83 | 4 | 0.91 (0.65, 1.26) | 102 | 8 | **0.28 (0.22, 0.36)** |
|  | ≥ 65 years | 560 | 102 | **0.55 (0.50, 0.61)** | 189 | 29 | **0.71 (0.59, 0.85)** | 199 | 41 | **0.36 (0.31, 0.42)** |

CHD, coronary heart disease.

Note: Statistically significant results are in bold.

### **Supplemental Table A7.** Meta-analysis of the association between low-dose ASA use with all-cause and common subtype dementia incidence, stratified by both sex and coronary heart disease

| **CHD** | **Sex** | **All-cause dementia** | | | **Alzheimer’s disease** | | | **Vascular dementia** | | |
| --- | --- | --- | --- | --- | --- | --- | --- | --- | --- | --- |
|  |  | **n_case_**  **UK Biobank** | **n_case_**  **ESTHER** | **Meta-analysis**  **HR (95% CI)** | **n_case_**  **UK Biobank** | **n_case_**  **ESTHER** | **Meta-analysis**  **HR (95% CI)** | **n_case_**  **UK Biobank** | **n_case_**  **ESTHER** | **Meta-analysis**  **HR (95% CI)** |
| No | Female | 2,324 | 192 | **1.11 (1.05, 1.17)** | 929 | 73 | 1.02 (0.93, 1.11) | 510 | 74 | **1.23 (1.10, 1.38)** |
|  | Male | 2,414 | 165 | 0.96 (0.91, 1.02) | 828 | 51 | 1.02 (0.92, 1.12) | 626 | 59 | **1.17 (1.06, 1.29)** |
| Yes | Female | 269 | 55 | **0.59 (0.51, 0.68)** | 102 | 19 | **0.57 (0.46, 0.71)** | 82 | 19 | **0.40 (0.31, 0.52)** |
|  | Male | 577 | 64 | **0.42 (0.38, 0.47)** | 170 | 14 | **0.82 (0.66, 1.03)** | 219 | 30 | **0.30 (0.26, 0.35)** |

Statistically significant results are in bold.
